# Static and dynamic noise abolish altruistic biases in Parkinson’s disease independent of deep brain stimulation

**DOI:** 10.64898/2026.10.08.26365079

**Authors:** Fabrice Hubschmid, Manuela Sellitto, Bahne Hendrik Bahners, Thomas Pirenne, Tobias Kalenscher, Alfons Schnitzler, Marijn Van Wingerden, Esther Florin

**Affiliations:** Institute of Clinical Neuroscience and Medical Psychology, Medical Faculty and University Hospital Düsseldorf, Heinrich Heine University Düsseldorf, Düsseldorf, Germany; Clinical Psychology, Institute of Experimental Psychology, Heinrich Heine University Düsseldorf, Düsseldorf, Germany; Comparative Psychology, Institute of Experimental Psychology, Heinrich Heine University Düsseldorf, Düsseldorf, Germany; Department of Brain and Behavioral Sciences, University of Pavia, Pavia, Italy; Department of Neurology, Center for Movement Disorders and Neuromodulation, Medical Faculty and University Hospital Düsseldorf, Heinrich Heine University Düsseldorf, Düsseldorf, Germany; Department of Computational Cognitive Science, Research Center for Cognitive Science and Artificial Intelligence, Tilburg School for Humanities and Digital Sciences, Tilburg University, Tilburg, the Netherlands

**Keywords:** Parkinson’s disease, social discounting, drift diffusion, dynamic noise, deep brain stimulation, altruism

## Abstract

There are known alterations in decision-making attributed to Parkinson’s disease. Yet little is known about how those alterations affect decisions in social contexts. We aimed to assess social decision-making in healthy human volunteers as well as people with Parkinson’s disease treated with deep brain stimulation of the subthalamic nucleus. Twenty healthy controls and twenty-two people with Parkinson’s in both the stimulation ON and OFF conditions took part in a social discounting task that measured the degree of generosity toward others as a function of social-emotional distance. Using a computational approach, we modelled mechanisms of social discounting across three different models that differed in how they mapped choice valuation onto observed decisions. People with Parkinson’s less often made generous choices than controls. Across all analyses, we found converging evidence that this effect was attributable to increased static and dynamic decision noise, i.e. inhibitory and attentional processes, but not changes in prosocial valuation. While stimulation sped up decisions, there was no effect of deep brain stimulation on social decision-making. Present results indicate that people with Parkinson’s are more prone to random processes such as increased impulsivity and attentional lapses during choices in social situations. Such changes attributed to the disease appear to evade clinical care with dopaminergic compensation and deep brain stimulation.

## Introduction

While Parkinson’s disease is clinically characterized by motor symptoms, it typically comes with other symptoms as well. There is evidence in people with Parkinson’s disease (PwP) of impaired reinforcement learning (Frank et al., 2004), decision-making under conflict (Cavanagh et al., 2011), effort-based decisions (Pagnier et al., 2024), as well as increased risk-taking and egoistic choice (Florin et al., 2013; Rosen et al., 2015). Such changes can have a tangible impact on everyday social life, i.e. marital interactions, social adaptation, and problems within the workplace (Schüpbach et al., 2006). These behavioural changes can even worsen with deep brain stimulation (DBS) of the subthalamic nucleus (STN) and dopaminergic medication (Frank et al., 2007; Mosley et al., 2020). While extensive prior research has examined certain aspects of decision-making in Parkinson’s disease, such as reinforcement learning (Frank et al., 2004; Tichelaar et al., 2023), it remains unclear whether and how the disease affects social decision-making.

Parkinson’s disease is characterized by changes in both the time needed to perform motor responses and changes in the regulation of decision times as a matter of conflict, i.e. response caution (Cavanagh et al., 2011; Herz et al., 2018; Low et al., 2002). This makes the assessment of behaviour in PwP vulnerable to speed-accuracy trade-offs if the time needed to decide is not accounted for. As such, it is also possible that changes in regulatory mechanisms of decisional conflict in PwP act on the conflict between self- and other-regarding motives in social situations (Kalenscher et al., 2025).

Further, changes in executive functions such as working memory and attention are prevalent non-motor symptoms of Parkinson’s disease (Biundo et al., 2025; Dujardin et al., 2013). Therefore, it is important to account for possible attentional lapses in behavioural sequences in PwP. Periods of attentional lapses or task disengagement may render behaviour more stochastic (Li et al., 2024). As human volunteers are prone to hyperaltruistic biases in experimental set-ups (Crockett et al., 2015), if PwP are more prone to lapses than healthy controls then revealed differences in prosociality in Parkinson’s may not reflect true social preferences but may be attributed to dynamic decision noise.

To account for all these prosocial decision-formation mechanisms we implemented different computational models of social discounting that differed in their policy i.e. how they mapped modelled internal choice preferences onto observed actions. As such our approach accounted for changes in the sampling of decision relevant information as a function of time as well as possible attentional lapses and the dynamic evolution of noise over time in an established social discounting paradigm. The task measured prosocial decision-making as a function of the social-emotional distance between the decision-maker and the recipient of the procosial act (Kalenscher et al., 2025; Sellitto et al., 2021; Sellitto & Kalenscher, 2022; Strombach et al., 2015). We investigated how those mechanisms operate in PwP implanted with STN-DBS, both ON and OFF stimulation, as well as age- and gender-matched controls in a between-subjects experiment.

## Materials & Methods

### Participants

The study was approved by the local ethics committee (Ethic Committee of the Medical Faculty of Heinrich Heine University Düsseldorf (2019-568-KFogU)), and data acquisition was done in accordance with the latest revised Declaration of Helsinki. Participants gave written informed consent before testing. PwP were recruited from the inpatient ward of the Center for Movement Disorders and Neuromodulation of University Hospital Düsseldorf.

To be included in the study, participants had to be older than 40 years. PwP additionally had to be diagnosed with Parkinson’s disease and be implanted with DBS electrodes in the STN for at least 6 months. Participants were excluded if they had a severe neurological, psychiatric, or internal disease, except for Parkinson’s disease.

The final sample consisted of 20 healthy participants (HC) (4 women, age: M = 64.65, SD = 9.31) as well as 22 PwP (3 women, age: M = 62.00, SD = 9.42). A more detailed characterisation of the study sample can be found in the supplementary materials (see supplementary materials, Section 1). PwP completed the task on dopaminergic therapy, which corresponds to their normal everyday state.

### Social discounting task

We used an established social discounting task that measures prosociality in relation to the social/emotional distance of the recipient of a generous act (Sellitto et al., 2021; Strombach et al., 2015).

Prior to the task, participants were asked to think about people from their social environment who would correspond to different social distances. Participants had to report first names as well as the nature of the relationship they shared with that person. The scale ranged from 1 (socially closest person) to 100 (complete stranger). As in previous versions of the task, they were asked to choose six existing persons for the social distances 1, 5, 10, 20, 50, and 100.

Every trial started with a display of the social distance information (Fig. 1A). The social distance scale was represented as a dotted line. Information about the person with whom the trial was to be played (the partner) was indicated with a yellow icon depending on the distance on the scale. The participant was represented with a purple icon at the left end of the scale. Information about the social distance was displayed for a jittering interval of 4 to 6 seconds.

**Figure 1.**
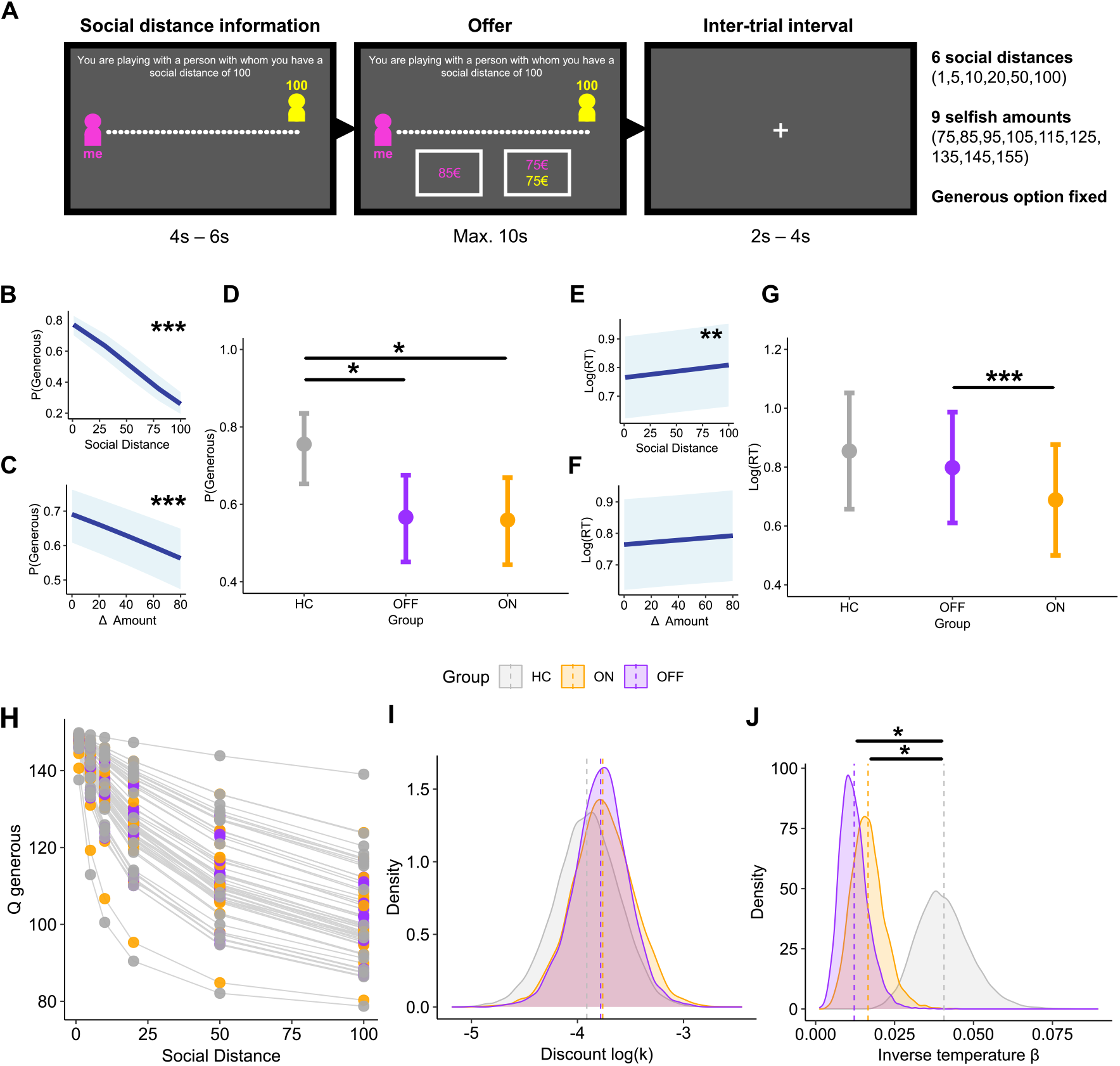
Task, inferential analyses, and social discounting. (**A)** Schematic depiction of the social decision-making task. **(B)** Estimated fixed effect of the social distance on the probability of generous choice from the generalized mixed logistic regression model (GLM). **(C)** Estimated (GLM) fixed effect of the amount difference on the probability of generous choice. **(D)** Estimated (GLM) probability of generous choice depending on group. **(E)** Estimated fixed effect of the social distance on the (log) reaction time (RT) from the linear mixed model (LMM). **(F)** Estimated (LMM) fixed effect of the amount difference (Δ Amount) on the (log) reaction time. **(G)** Estimated (LMM) (log) reaction time depending on group. **(H)** Modelled discount in value of the generous choice options for one selfish amount (85) as example. Posterior distributions of the log (*k*) (**I**) and *β* (**J**) hyperparameters of the *SoftMax-Discount* model. In plots B, C, E, and F, shaded blue areas represent the 95% confidence interval. In plots E and H, the error bars represent the 95% confidence interval. Significance marked as: “***” *p* < 0.001, “**” *p* < 0.01, “*” *p* < 0.05. In plots showing distributions of hyperparameters (I, J) significance is drawn from comparisons through difference densities.

In the choice interval, participants chose between a generous and a selfish option. The generous choice option was fixed and would always lead to both the participant and the partner in the trial receiving 75€. In the selfish option, the amount the participant would receive was always equal or higher than in the generous option. Participants were instructed that selection of the selfish choice would result in the partner not receiving anything. Nine different selfish amounts were used in the task (75€, 85€, 95€, 105€, 115€, 125€, 135€, 145€, 155€). Participants had a maximum of 10 seconds to choose using the left and right arrow keys of the keyboard. The choice period was followed by an inter-trial-interval that jittered between 2 and 4 seconds.

The position of the choice options on the screen and their corresponding buttons were randomized and counterbalanced. The full trial sequence consisted of all combinations of social distances and selfish amounts repeated twice in both choice positions on the screen for a total of 108 trials. Selected amounts were not paid out at the end of the testing session, but we used an incentivization procedure to create a monetary and social incentive to complete the task. Namely, one of the trials was randomly selected, and the amount selected was then paid out to either the participant or to the participant and the corresponding partner in the selected trial.

PwP completed the task twice on the same day. Half of the PwP first completed the experiment in the stimulation ON state, and the other half in the stimulation OFF state. The task was completed on a portable computer, and the keyboard was used as the response unit.

### Model agnostic analyses

Data analysis was done in R (*v*.*4*.*3*.*3*: R Core Team, 2022) and Stan (*v*.*2*.*32*.*2*: Carpenter et al., 2017). Visualisations were generated using *ggplot2* (Wickham, 2011). Trials with no responses were excluded.

To assess the effect of the social distance, the difference in pay-outs between choices (amount difference), and the group (HC, ON vs. OFF) on the probability of a generous choice, we fitted a mixed logistic regression model, while for the (log) reaction times, a linear mixed model was used. Models considered the social distance and amount difference (i.e. the difference in payouts for the decision-maker between the generous and selfish choices) as well as the group as fixed effects. The participant was added as a random variable and only random intercepts were modelled. To test for differences between groups, we conducted post-hoc comparisons of marginal means corrected for multiplicity through Tukey adjustments (see supplementary materials, Section 2). Models were fitted to the data using the *lme4* (Bates et al., 2015) and *lmertest* (Kuznetsova et al., 2017) software packages.

Given that the PwP were tested in the medicated state, as a control analysis we additionally tested for possible effects of the levodopa equivalent daily dose on prosocial behaviour and reaction times in the task (see supplementary materials, Section 3).

### Computational modelling of social discounting

We characterized the mechanisms underlying observed choices using computational modelling. We implemented a modelling strategy that aimed at: **1**. Characterize the rate of hyperbolic discounting across social distance, **2**. Account for possible changes in the regulation of decision-times as a matter of conflict in PwP, and **3**. Account for possible effects of attention and concentration on task performance. This was done through the implementation of three different variants of a social discounting model that only differed in their policies i.e. how they mapped internal value preferences to observed choices. Models were implemented in a hierarchical Bayesian framework (f.e. Ahn et al., 2017; Piray et al., 2019; Wiecki et al., 2013).

The data from one PwP was excluded from computational analyses because this participant chose the generous choice option only once in the stimulation ON state. The data from two healthy controls, who only chose the generous option in the task, were also excluded. We removed their data to prevent extreme behavioural patterns from impairing estimation of latent parameters fitted on the probabilistic occurrence of choices and to preserve generalizability of group level comparisons.

#### Assessment of social discounting

First, we modelled observed choice probabilities using social discounting as it is traditionally determined:

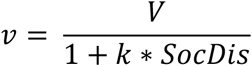

Where the valuation of generosity *ν* is hyperbolically discounted as a function of the social distance *SocDis* determined by slope *k* and intercept *V*. This discount is known to be hyperbolic rather than exponential (Jones & Rachlin, 2006). This phenomenon has been replicated across numerous studies and cohorts (Kalenscher et al., 2025; Rhoads et al., 2023; Sellitto et al., 2021; Sellitto & Kalenscher, 2022; Strombach et al., 2015). In practice, when fitting to observed choices rather than indifference points, the rate of hyperbolic discounting *k* is estimated as (Sellitto et al., 2021):

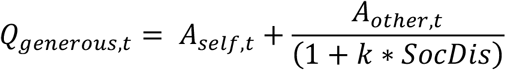

Where the subjective value given to the generous choice option *Q*_*generous,t*_ in a trial is determined by the amount of money that can be obtained by the decision maker him/herself *A*_*self,t*_ as well as the amount of money that can be obtained by the partner in the trial *A*_*other,t*_. The value of the payout of others is still hyperbolically discounted as a function of the social distance of the partner in the trial *SocDis*.

As in the selfish choice, the payout is only for the decision-maker themselves *SocDis =* 0, the amount obtained upon selection of the selfish choice solely determines the value of the selfish choice *Q*_*selfish,t*_ .

The probability of selecting the generous choice option in a trial *P*_*generous,t*_is then estimated through a SoftMax policy:

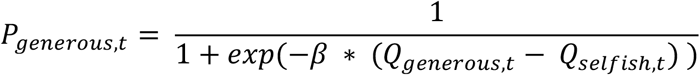

Where *β* is the inverse decision temperature that scales how deterministically value differences drive action selection.

This model (Model1: *SoftMax-Discount*) was used as a baseline for the assessment of social discounting in the task and corresponds to a hierarchical Bayesian implementation of the model used in Sellitto et al. (2021). As is traditionally done with discount rates, we report them in log-space (f.e. Kalenscher et al., 2025; Peters & D’Esposito, 2020).

#### Discount drift diffusion modelling

In a second step, we modelled choice-reaction time distributions with a drift diffusion model variant (Ratcliff et al., 2016; Ratcliff & McKoon, 2008; Wiecki et al., 2013). For these analyses, we additionally removed trials with outlying reaction times, as they are likely caused by processes not considered in the DDM (Wiecki et al., 2013). We defined outliers as the extreme 5% of trials and removed them from the upper and lower tails of the reaction time distribution before model fitting.

The DDM describes an accumulation to bound process where a decision variable stochastically approaches one of two decision boundaries. Formally, it models the probability of one choice over the other and the associated reaction time through a Wiener-first-passage-time (*wfpt*) distribution of four free parameters.

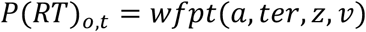

The boundary separation *a* determines the distance between the two boundaries and is usually interpreted in terms of response caution. The non-decision time *ter* corresponds to the time not allocated to accumulation of decision-relevant evidence. The starting point bias *z* determines where between the lower and upper boundary evidence accumulation starts. The drift rate *ν* is the average rate at which evidence is stochastically approaching one boundary rather than the other.

Previous work in the field of reinforcement learning has suggested using the DDM to map modelled values to observed choices effectively using it as a policy rather than a sole process model (Pedersen et al., 2017; Pedersen & Frank, 2020). This idea has been extended to discounting experiments by Peters and D’Esposito (2020).

In these models, the drift rate *ν*_*t*_ varies trial-by-trial and is influenced by the difference between the value given to both choices. The trial drift rate *ν*_*t*_ is determined by the scaled value difference between the two choices. These models include an additional free parameter, the drift rate coefficient *ν*_*coeff*_ that scales the subjective value difference between choices.

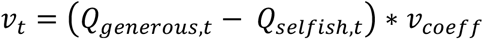

This corresponds to the so-called linear mapping of value differences onto a drift diffusion process (Pedersen et al., 2017; Pedersen & Frank, 2020).

Choice probabilities and associated reaction times in a trial are then estimated from a DDM with a fixed bias term so that:

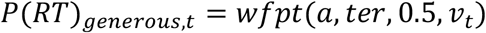

In this model (Model 2: *DDM-Discount*), we decided to fix the bias term to the non-biasing value of 0.5. This was done because including it as a free parameter created trade-offs in explained variance between *z* and *ν*_*coeff*_. We decided against a parametrization of the model with the sigmoidal mapping of value differences (Fontanesi et al., 2019), because the inclusion of the drift rate asymptote parameter impaired recovery (see supplementary material, Section 4)

#### Hidden Markov modelling of dynamic noise

Both the SoftMax and the DDM policies assume static levels of noise over the trial sequence. However, noise can be caused by different latent processes such as exploration-exploitation dynamics (Chakroun et al., 2020; Daw et al., 2006; Wilson et al., 2014), speed accuracy trade-offs (Ratcliff et al., 2016; Voss et al., 2004), but also attention and concentration (Esterman & Rothlein, 2019). Depending on the cause, the assumption that noise and policies remain static over a task with multiple trials can be unrealistic. In periods where participants disengage from the task, noise levels tend to be temporally autocorrelated (Li et al., 2024; Venditto et al., 2024). This is particularly relevant when assessing decision-making in neurological cohorts such as PwP, where attention and concentration are expected to change (Biundo et al., 2025; Dujardin et al., 2013).

To account for the temporal evolution of noise levels, in the last model (Model 3: *HMM-Discount*) we used the dynamic noise estimation suggested and described by Li et al. (2024). Broadly, this method makes use of a Hidden Markov model (HMM) to estimate occupancy of periods of task engagement (engaged state) and attentional lapses (random state) from behavioural sequences.

In this model, the likelihood of selecting the generous choice option is given by a weighted mixture of observing the generous choice in the engaged and random states.

The probability of observing a generous choice in the engaged state in a trial *P*_*engaged,t*_ is determined by the hyperbolic value function and SoftMax action selection as described above. In the random state the probability of observing any choice is *P*_*random,t*_ = 0.5. In any given trial, the probability of making a generous choice *P*_*generous,t*_ is estimated from:

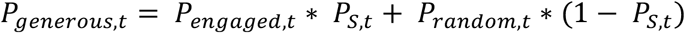

Where *P*_*s,t*_ corresponds to the probability of being in the engaged state i.e. actively playing the task and complying with the expected strategy, and its inverse 1 − *P*_*s,t*_ ,the probability of being in the random state. At the beginning of the trial sequence *P*_*s,t*_ is initialized at 0.5 and after every trial *P*_*s,t*_ is updated according to the transition probabilities of the HMM:

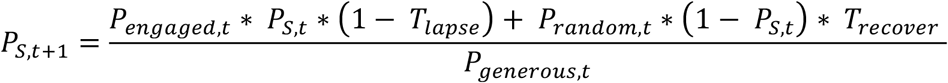

Where *T*_*lapse*_ and *T*_*recover*_ are free parameters that determine the transition probability of switching from the engaged to the random state (*T*_*lapse*_) and the transition probability of switching from the random to the engaged state (*T*_*recover*_). In this model we fixed the inverse SoftMax temperature *β* to 0.4. This was done to help parameter identifiability, as static noise is a specific case of dynamic noise (Li et al., 2024). In order for the model to correspond to the original dynamic noise implementation of Li et al., (2024), it was implemented in a non-hierarchical way.

#### Model fitting and posterior inference

Models were implemented in a hierarchical Bayesian framework using non-centred parametrizations (f.e. Ahn et al., 2017; Wiecki et al., 2013). Posterior parameters were estimated using no-U-turn Markov Chain Monte Carlo sampling (NUTS-MCMC) with the *rstan* software package (Stan Development Team, 2024). For group- and subject-level parameters we used the weakly informative *normal*(0,1) priors with the exception of DDM hyperparameters for which literature informed priors were used (Matzke & Wagenmakers, 2009). For standard deviation parameters we used *normal*(0,02).

Posterior parameter distributions were sampled from four independent chains. All chains ran for 4000 iterations, with the first 2000 being burned out. The chains were concatenated at the end for a total of 8000 draws used for posterior inference. Convergence of chains was assessed through 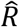 , with convergence considered adequate if all values < 1.1 (Gelman & Rubin, 1992). The fitting procedure was repeated independently in all groups (HC, PwP ON STN-DBS, and PwP OFF STN-DBS).

To compare mechanisms of social decision-making between groups, we used the posterior distributions of (group) hyperparameters. To this end, we calculated difference densities by subtracting posterior draws of one parameter from another. A significant difference between two parameters was assumed if the 95% highest density interval of the difference density (HDI-DD) did not overlap with 0. Plots corresponding to each posterior comparison are provided in the supplementary materials (see supplementary materials, Section 5). For the *HMM-discount* model which was non-hierarchical, we compared individual level parameters using independent and dependent non-directed Bayesian t-tests as implemented in JASP (*v*.*0*.*19*.3: Love et al., 2019). Finally, to assess group differences in the probability of task engagement we used a linear mixed model. The model used the estimated posterior probability of being in the engaged state *P*_*s,t*_ in every trial of the task as dependent variable. The group (3 level factor) was used as fixed effect, and the participant was included as random effect. Only random intercepts were modelled.

To ensure the fitted models were able to predict the empirical data in meaningful ways, and that the parameters could be properly identified from the task, we ran posterior predictive checks and parameter recovery analyses (Wilson & Collins, 2019). We report those validation analyses in the supplementary materials (see supplementary materials, Section 4).

## Results

### PwP were less generous than controls

Both the increase in social distance (fixed effect, coefficient = -0.023 [-0.025, -0.021], p < 0.001) and in the amount difference (fixed effect, coefficient = -0.007 [-0.009, - 0.005], p < 0.001) significantly decreased the probability of generous choice (Fig. 1B-C). This suggests the task induced the expected social discounting behaviour. Compared to the control group, the probability of selecting generous options was lower in PwP both ON (fixed effect, coefficient = -0.888 [-1.598, -0.199], p < 0.01) and OFF DBS (fixed effect, coefficient = -0.858 [-1.568, -0.170], p < 0.013). Post-hoc comparisons indicated no significant differences between PwP ON and OFF stimulation (Fig. 1D; see supplementary materials, Section 1).

The social distance also had a significant effect on the reaction time in the task (fixed effect, coefficient = 0.0004 [0.0001, 0.007], p < 0.008), i.e. participants’ response times increased as a function of the social distance possibly indicating increased conflict (Fig. 1E-F). Post-hoc comparisons indicated that reaction times were shorter in PwP ON DBS (M = 0.704, SD = 0.687) compared to OFF DBS (M = 0.795, SD = 0.723; p < 0.001) (Fig. 1G). For reaction times, no comparison between HC and PwP was significant.

### Value differences influenced action selection less deterministically in PwP compared to controls

Posterior comparisons from the *SoftMax-Discount* model showed no differences in the rate of hyperbolic discounting log(*k*), between groups (Fig. 1H-I). However, the inverse decision temperatures *β* were significantly decreased in PwP compared to the control group (HC vs ON: HDI-DD: lower = 0.052, upper = 0.044; HC vs OFF: lower = 0.01, upper = 0.048) (Fig. 1J).

Similarly, posterior comparisons from the *DDM-Discount* model (Fig. 2A) also showed no differences in the rate of discounting log(*k*) (Fig. 2B). However, the non-decision times were significantly shorter in the stimulation ON and OFF group, separately, compared to the HC group (HC vs ON: HDI-DD: lower = 0.277, upper = 0.443; HC vs OFF: lower = 0.171, upper = 0.358). This is a known effect in PwP (Gupta et al., 2022). DBS-ON, compared to OFF, further shortened non-decision time (HDI-DD: lower = 0.002, upper = 0.198) (Fig. 2C).

**Figure 2.**
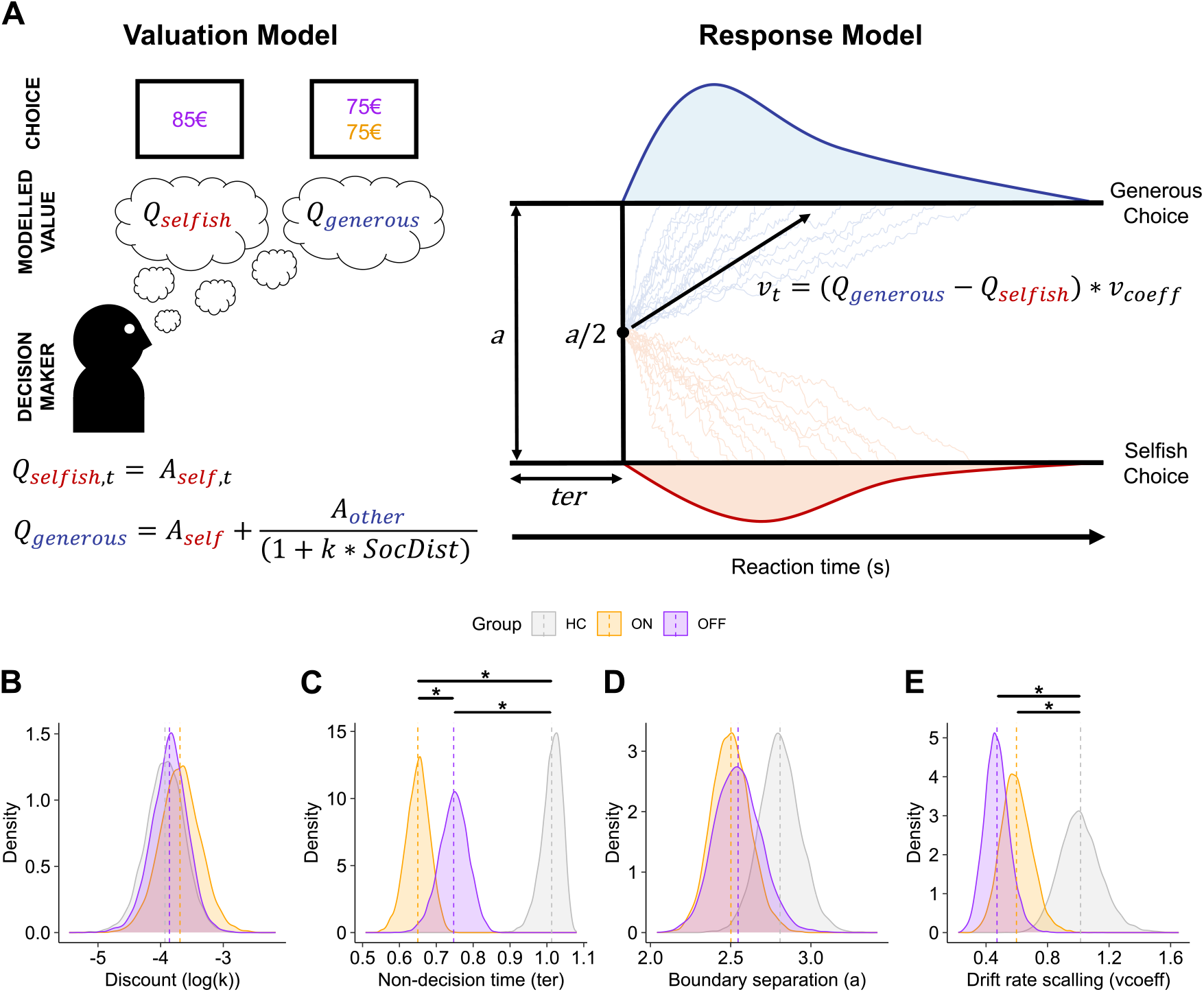
Modelling of choice-reaction time distributions. **(A)** Schematic representation of the *DDM-Discount* model. The model combines a value function and a response model. The valuation model assumes that the subjective (*Q*) value of the generous choice is hyperbolically discounted based on the social distance. The response model (policy) is a drift diffusion model with a fixed bias term, where the drift rate is allowed to vary trial-by-trial based on differences in expected (*Q*) values. Value differences are scaled by the drift rate coefficient *ν*_*coeff*_ which determines how deterministically the expected value difference is driving the drift rate upwards or downwards towards the favoured choice. (**B**) Posterior distributions of the discount log (*k*) hyperparameter. (**C**) Posterior distributions of the non-decision time *ter* hyperparameter. (**D**) Posterior distributions of the boundary separation *a* hyperparameter. (**E**) Posterior distributions of the drift rate coefficient (*ν*_*coeff*_) hyperparameter. In plots showing distributions of hyperparameters significance is drawn from comparisons through difference densities.

There was a substantial but non-significant decrease in boundary separations in PwP compared to the HC group both in the ON and OFF stimulation state (HC vs ON: HDI-DD: lower = -0.037, upper = 0.661; HC vs OFF: lower = -0.11, upper = 0.641).

To assess how deterministically prosocial evidence accumulated over time, we tested for differences in the drift rate coefficient *ν*_*coeff*_. Both the difference between the HC and the stimulation ON groups (HDI-DD; lower = 0.081, upper = 0.744) and between the HC and the stimulation OFF groups (HDI-DD; lower = 0.232, upper = 0.851) were significant. Therefore, regardless of the DBS state, differences in values less deterministically influenced prosocial evidence accumulation in PwP than HC (Fig. 2D).

#### PwP spent more trials in the random state than controls

Bayesian independent samples t-tests revealed that the probability of recovering from an attentional lapse *T*_*recover*_ was lower in PwP OFF STN-DBS compared to controls with strong evidence for the alternative hypothesis (BF_10_ = 11.718) and ON STN-DBS compared to controls with weak evidence for the alternative hypothesis (BF_10_ = 2.709) (Fig. 3A-E).

**Figure 3.**
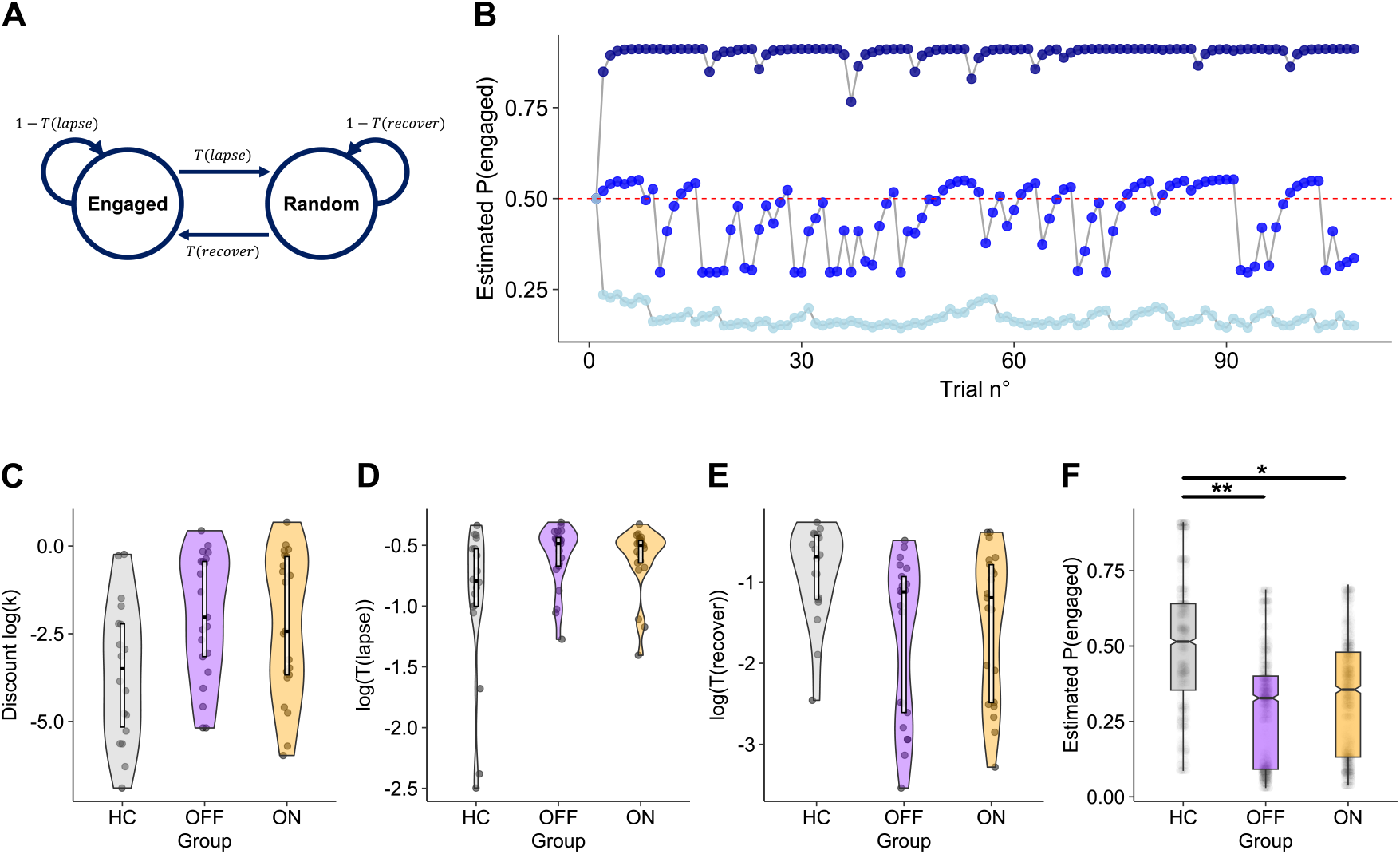
Hidden Markov modelling of dynamic noise. **(A)** Schematic representation of the hidden Markov model (HMM) for dynamic noise estimation of Li et al., (2024). The model estimates choice probabilities from a two-state HMM that assumes decision makers are either in a random or an engaged state. Transition from the engaged to the random state is determined by the probability of lapsing (*T*_*lapse*_). Transition from the random to the engaged state is determined by the probability of recovering (*T*_*recover*_). (**B**) Model estimated posterior probability of task engagement over the trial sequence for three exemplary participants: One participant that followed the expected discounting strategy throughout the entire task (darkblue), one participant that responded randomly throughout the task (lightblue), and one participant that switched between periods of task engagement and attentional lapses (blue). (**C**) Individual-level parameters for the discount log(*k*) (**D**), the probability of lapsing log (*T*_*lapse*_) (**E**) and the probability of recovering log (*T*_*recover*_). (**F**) Model estimated posterior probability of being in the engaged state in every trial per group. For the boxplot and the boxes within the violin, the boxes represent the median and the interquartile range. Significance in plot F comes from a linear mixed model assessing the fixed effect of the group on the probability of task engagement. Significance marked as: “**” *p* < 0.01, “*” *p* < 0.05.

The probability of lapsing *T*_*lapse*_ was also generally increased in PwP compared to HC. The evidence for a difference was however only weak in both comparisons (ON vs HC: BF_10_ = 1.833; OFF vs HC: BF_10_ = 2.638) (Fig. 3D).

The linear mixed model revealed a significant effect of the group on the model estimated posterior probability of being in the engaged state in a trial *P*_*s,t*_. Compared to the control group, the probability of being in the engaged state was lower in PwP both ON (fixed effect, coefficient = -0.179 [-0.308, -0.050], p = 0.01 and OFF STN-DBS (fixed effect, coefficient = -0.213 [-0.342, -0.084], p = 0.003) (Fig. 3F).

## Discussion

In the present study, we assessed mechanisms of prosocial decision-making of PwP. We observed a decrease in generous choice in Parkinson’s disease, which aligns with existing evidence (Rosen et al., 2015). Across all models we found that this reduction in generosity can be attributed to parameters that determine how stochastically participants behave but not to differences in how participants valued generous choices i.e. prosociality.

To disentangle contributions of choice valuation and sampling of evidence in time, we used a model that mapped value differences onto a DDM. Such models have grown in popularity in recent years (f.e. Berwian et al., 2020; Fontanesi et al., 2019; Hubschmid et al., 2025; Pedersen et al., 2017; Pedersen & Frank, 2020; Reimer et al., 2024)) and have already proven useful to interpret behavior of neurological patients (Peters & D’Esposito, 2020). They allow the concomitant assessment of choice valuation, accumulation of evidence in time, and mechanisms that lead to the resolution of conflict, all known to be impacted by Parkinson’s disease (Frank et al., 2007; Herz et al., 2018; Pagnier et al., 2024; Tichelaar et al., 2023). In this model, both the drift coefficient and the boundary separation decreased for PwP in our study. This indicates that choices in social contexts are more prone to random influences and impulsivity rather than reflecting true preferences for selfish choices in PwP. While HC show a clear preference for generous choices in this design, PwP are more likely to show more random choices and thus decreased generosity under these conditions.

Connectivity from the STN to prefrontal areas is thought to be involved in the conflict between habitual and controlled responses (Frank et al., 2007; Herz et al., 2024). The observed decrease in use of choice valuation and response caution in PwP possibly indicates that fronto-basal connectivity acts as a less efficient “brake signal” (Cavanagh et al., 2011). This is possibly reflected by an earlier termination of evidence accumulation and initiation of choice within pre- and motor areas (Gupta et al., 2022). This could decrease the importance of prefrontal choice valuation, leading to less deliberate choices in PwP.

To account for temporal changes in decision-making noise, we used the dynamic noise estimation method of Li et al. (2024). Those analyses showed that PwP mostly differed from HC in how likely they were to recover from periods of task disengagement. Consequently, PwP spent longer periods of the task in the random state, which further contributed to abolishing the bias for generous choices observed in healthy individuals. This emphasizes the relevance of dynamic noise estimation in the study of behaviour in Parkinson’s disease, as failure to account for periods where participants are not complying with expected task strategies could contribute to erroneous conclusions regarding cognitive and behavioural impairments in PwP.

Across all analyses, we found no effect of STN-DBS on prosocial decision-making, in addition to speeding motor processes, i.e., faster reaction times and shorter non-decision times ON DBS compared to OFF DBS. We also tested for an association between model parameters and the distance between the DBS electrodes and the motor and associative areas of the STN and found no relationship (see supplementary materials, Section 6). Present results suggest the absence of an effect of STN-DBS on social discounting. This aligns with previous evidence that shows that temporal discounting is not modulated by STN-DBS (Seinstra et al., 2016; Seymour et al., 2016) and possibly indicates a general absence of DBS effect on discounting mechanisms across domains.

An important limitation of our study is that PwP were tested under dopaminergic therapy, as this reflects the “normal” everyday state. However, this could be a confound because intake of levodopa has been associated with decreased altruistic behaviour (Crockett et al., 2015). Further, PwP show increased motivation to complete effortful prosocial choices under dopaminergic medication (Talbot et al., 2026), making it inconclusive how dopamine influences prosocial choices in PwP. Future studies should manipulate the medication state in addition to DBS. Such a two-factorial design has already yielded novel insights into decision-making in Parkinson’s disease (Pagnier et al., 2024).

In sum, we investigated prosocial decision-making in PwP implanted with STN-DBS and healthy controls. We observed a decrease in generous choice in Parkinson’s disease, which aligns with existing evidence (Rosen et al., 2015). With converging evidence across all analyses, we found that decisions in Parkinson’s disease were more prone to stochastic influences, abolishing biases towards generous choices observed in HC leading to more selfish choices, despite not necessarily reflecting true preferences. Importantly, such behavioural changes appear to evade standard clinical care with dopaminergic compensation and deep brain stimulation.

## Data availability

Anonymized task data needed for replication of the present analyses will be made publicly available on a dedicated OSF repository upon acceptance of the present manuscript for publication.

## Code availability

Software scripts used for the replication of the results and plots reported in the main text are openly available on a dedicated GitHub repository (https://github.com/HubschmidF/Social_Decision_Making_in_PD). Any additional information potentially required for the re-analysis of the data can be obtained upon request to the corresponding author.

## Acknowledgments

We thank Iris Reis for data acquisition and Simon Desch for feedback on the analysis. We thank Tim Angelike for help in the stan implementation of dynamic noise modelling. BHB acknowledges support from the Thiemann Foundation (Parkinson Fellowship 2022). This study was supported by the Volkswagen Foundation (Lichtenberg program 89387) and the Deutsche Forschungsgemeinschaft (DFG, German Research Foundation) – Project-ID 424778381 – TRR 295.

## Author contributions

**F.H**: Data curation, formal analysis, visualization, software, writing manuscript. **M.S**: Conceptualization, data curation, editing the manuscript. **B.H.B**: Data curation, editing the manuscript. **T.P**: Formal analysis, editing manuscript. **T.K**: Conceptualization, editing the manuscript. **A.S**: Conceptualization, editing the manuscript. **M.V.W**: Conceptualization, editing the manuscript. **E.F**: Supervision, data curation, conceptualization, editing manuscript.

