## Supplementary_Materials for "Static and dynamic noise abolish altruistic biases in Parkinson’s disease independent of deep brain stimulation"

Social discounting in Parkinson's


Fabrice Hubschmid [1,2], Manuela Sellitto [3,4], Bahne Hendrik Bahners [1,5], Thomas Pirenne [1], Tobias Kalenscher [3], Alfons Schnitzler [1,5], Marijn Van Wingerden [6] & Esther Florin [1]

*Affiliations*

[1] Institute of Clinical Neuroscience and Medical Psychology, Medical Faculty and University Hospital Düsseldorf, Heinrich Heine University Düsseldorf, Düsseldorf, Germany

[2] Clinical Psychology, Institute of Experimental Psychology, Heinrich Heine University Düsseldorf, Düsseldorf, Germany

[3] Comparative Psychology, Institute of Experimental Psychology, Heinrich Heine University Düsseldorf, Düsseldorf, Germany

[4] Department of Brain and Behavioral Sciences, University of Pavia, Pavia, Italy

[5] Department of Neurology, Center for Movement Disorders and Neuromodulation, Medical Faculty and University Hospital Düsseldorf, Heinrich Heine University Düsseldorf, Düsseldorf, Germany

[6] Department of Computational Cognitive Science, Research Center for Cognitive Science and Artificial Intelligence, Tilburg School for Humanities and Digital Sciences, Tilburg University, Tilburg, the Netherlands


***Sections***


*Section 1*: Participant characteristics

*Section 2*: Tukey corrected post-hoc comparisons for mixed effects models

*Section 3*: Control analyses for levodopa dosage

*Section 4:* Posterior predictive checks and parameter recovery simulations

*Section 5:* Inference on posterior hyper-parameters

*Section 6*: Correlations between electrode localizations and model parameters

Supplemental References

### Section 1

#### Participant characteristics

**Supplementary table 1: Participant characteristics**

| | People with Parkinson's Disease | | HC | $BF_{10}$ ON≠OFF | $BF_{10}$ PwP≠HC |
|---|---|---|---|---|---|
| | DBS-ON | DBS-OFF | | | |
| Counts (n) | 22 | - | 20 | - | - |
| Age | 62.00 (9.42) | - | 64.65 (9.31) | - | 0.424 |
| Gender (f/m) | 3/19 | - | 4/16 | - | - |
| UPDRS-III | 25.00 (15.67) | 35.05 (15.41) | - | > 100 | - |
| MMSE | 28.45 (2.09) | - | 29.45 (0.89) | - | 1.394 |
| SES-17 | 11.55 (2.96) | - | 10.5 (2.91) | - | 0.515 |
| BDI-II | 14.05 (9.48) | - | 5.5 (5.71) | - | 27.275 |

**Note.** To compare groups, we used paired and independent samples Bayesian t-tests to calculate Bayes Factor. The Bayes Factor ($BF_{10}$) is an indication of how much the data provides evidence for a difference or similarity between groups. UPDRS-III = Unified Parkinson's Disease Rating Scale part-III; MMSE = Mini Mental State Examination; SES-17 = German version of the Social Desirability Scale-17; BDI-II; Beck Depression Inventory. UPDRS-III data from one participant is missing.

### Section 2

#### Tukey corrected post-hoc comparisons for mixed effects models

**Supplementary table 2: Contrast comparisons for the choice probability GLM**

| Contrast | Estimate | SE | p-value |
|---|---|---|---|
| HC – OFF | 0.858 | 0.346 | **0.035** |
| HC – ON | 0.887 | 0.346 | **0.027** |
| OFF – ON | 0.029 | 0.067 | 0.898 |

**Note.** Pairwise comparisons of marginal means between groups. Results are averaged over the levels of amount difference and social distance. p-values were corrected through Tukey adjustments. (SE = Standard Error).

**Supplementary table 3: Contrast comparisons for the reaction time LMM**

| Contrast | Estimate | SE | p-value |
|---|---|---|---|
| HC – OFF | 0.056 | 0.139 | 0.914 |
| HC – ON | 0.166 | 0.139 | 0.457 |
| OFF – ON | 0.110 | 0.014 | **< 0.001** |

**Note.** Pairwise comparisons of marginal means between groups. Results are averaged over the levels of amount difference and social distance. p-values were corrected through Tukey adjustments. (SE = Standard Error).

#### ***Section 3***

**Control analyses for levodopa dosage**

Dopamine is well known to act on reward processing (Schultz et al., 1997). In the present study, we measured people with Parkinson's (PwP) in the normal, medicated state. However, it is possible that the tonic increase in dopamine due to medication acts on impulsivity and motivational behaviour (Frank et al., 2007). Further, the administration of levodopa has also been associated with reduced altruism in a social decision-making task (Crockett et al., 2015). Therefore, as a control analysis, we assessed whether the levodopa equivalent daily dose (LEDD) in PwP had an effect on both the probability of generous choice selection and the reaction time using mixed effect models.

To evaluate the effect of the LEDD on the probability of generous choice selection, we used a logistic regression model similar to that used for the main analyses, but only considering the data from PwP. This model considered the probability of generous choice selection as the dependent variable. To assess the effect of LEDD on the (log-transformed) reaction times, we used a similar linear mixed model as for the main analyses. Both models considered the group (stim ON, stim OFF), the amount difference, the social distance, and the LEDD as fixed effects. In both models, the participant was added as a random effect. Models were fitted to the data using the *lme4* (Bates et al., 2015) and *lmertest* (Kuznetsova et al., 2017) software packages. One participant was excluded from those analyses because of missing LEDD data.

Additionally, we assessed the necessity of including the LEDD as covariate (fixed effect) in the model using the Bayesian information criterion (BIC). Namely, we compared versions of the model as reported in the main text against a similar model additionally including the LEDD as fixed effect. The best-fitting model is chosen based on the lowest BIC.

The LEDD did not significantly influence the probability of generous choice (fixed effect, coefficient = -0.0006 [-0.002, 0.001], P = 0.465). The LEDD also did not have a significant effect on the (log) reaction time (fixed effect, coefficient = -0.0007 [-0.002, 0.0007], P = 0.357).

For the (log) reaction time, the model not including the LEDD as a predictor (BIC = 6941.4) was preferred compared to the model that included it (BIC = 6947.7). For the probability of generous choice, the model not including the LEDD (BIC = 5402.5) was also preferred compared to a model that included it (BIC = 5410.4).

#### ***Section 4***

**Posterior predictive checks and parameter recovery simulations**

In order to ensure the experiment was powered enough to provide conclusive evidence for the computational hypotheses tested and to assess whether the fitted models are able to predict empirical data in a meaningful way, we conducted parameter recovery analyses and posterior predictive checks (PPCs) (Palminteri et al., 2017; Wilson & Collins, 2019). For computational efficiency, we ran those analyses based on the parameter values of the healthy sample because, after the exclusion of the two excessively generous HCs from modelling analyses this was the smallest sample (n=18) and thus the least likely to benefit from group level shrinkage from the hierarchical implementation in parameter recoverability.

To this end we used Stan's (Carpenter et al., 2017) forward sampling tools to simulate synthetic datasets based on the parameter values of each posteriors NUTS-MCMC draw during fitting. This resulted in a total of 8000 synthetic datasets that were used for PPCs for each model. To assess the predictive accuracy of each model we plotted the choice that was the most likely to be predicted across all simulations and visually compared it to the empirically observed one. At the group level, we assessed how well models were able to predict the proportion of selfish and generous choices for every trial of the trial sequence. We found that models were all able to predict observed choices well above chance levels and that the models were very similar in predictive accuracy.

For each model, 10 synthetic datasets were randomly selected for further parameter recovery analyses. Models were then fitted again to the synthetic datasets with similar fitting routines as explained in the main text. We then assessed recovery of parameters by plotting true (generating) values against recovered ones. We considered recovery to be acceptable if the correlation between true and recovered values > 0.5 (f.e. Mancini et al., 2025; Wilson & Collins, 2019). In the implementation stages, we removed from consideration one version of the *DDM-discount model* that used an additional sigmoidal transformation on value differences because it impaired recovery (Fontanesi et al., 2019) (See Supplementary table 5).

**Supplementary table 4: Subject-level predictive accuracy**

| Participant | Accuracy *SM Discount* | Accuracy *DDM Discount* | Accuracy *HMM Discount* |
|---|---|---|---|
| n°1 | 0.692 | 0.692 | 0.692 |
| n°2 | 0.692 | 0.682 | 0.71 |
| n°3 | 0.491 | 0.472 | 0.519 |
| n°4 | 0.750 | 0.741 | 0.769 |
| n°5 | 0.796 | 0.787 | 0.806 |
| n°6 | 0.694 | 0.713 | 0.704 |
| n°7 | 0.685 | 0.694 | 0.704 |
| n°8 | 0.794 | 0.794 | 0.794 |
| n°9 | 0.611 | 0.620 | 0.630 |
| n°10 | 0.667 | 0.685 | 0.704 |
| n°11 | 0.787 | 0.787 | 0.796 |
| n°12 | 0.907 | 0.870 | 0.907 |
| n°13 | 0.907 | 0.907 | 0.907 |
| n°14 | 0.537 | 0.546 | 0.565 |
| n°15 | 0.829 | 0.838 | 0.867 |
| n°16 | 0.632 | 0.632 | 0.632 |
| n°17 | 0.830 | 0.830 | 0.830 |
| n°18 | 0.750 | 0.759 | 0.778 |
| **Average** | **0.725** | **0.725** | **0.740** |

**Note.** Posterior predictive checks. Proportions of accurately predicted choices in each participant for each model. (SM = SoftMax, DDM = Drift diffusion model, HMM = Hidden Markov model).

**Supplementary table 5: Parameter recovery**

| Model | Parameter | $r$ | $rho$ |
|---|---|---|---|
| *SM Discount* | $k$ | 0.873 | 0.751 |
| *SM Discount* | $\beta$ | 0.848 | 0.791 |
| *DDM Discount* | $k$ | 0.717 | 0.762 |
| *DDM Discount* | $a$ | 0.895 | 0.820 |
| *DDM Discount* | $ter$ | 0.972 | 0.908 |
| *DDM Discount* | $v_{coeff}$ | 0.841 | 0.753 |
| *DDM Discount (Sigmoidal)* | $k$ | 0.791 | 0.794 |
| *DDM Discount (Sigmoidal)* | $a$ | 0.894 | 0.815 |
| *DDM Discount (Sigmoidal)* | $ter$ | 0.964 | 0.878 |
| *DDM Discount (Sigmoidal)* | $v_{coeff}$ | 0.251 | 0.268 |
| *DDM Discount (Sigmoidal)* | $v_{max}$ | 0.356 | 0.416 |
| *HMM Discount* | $\log(k)$ | 0.605 | 0.652 |
| *HMM Discount* | $\log(T_{lapse})$ | 0.714 | 0.610 |
| *HMM Discount* | $\log(T_{recover})$ | 0.522 | 0.464 |

**Note.** Parameter recovery. Correlations between true (generating) and recovered parameter values for all models. (r = Pearson's r, rho = Spearman's rho, SM = SoftMax, DDM = Drift Diffusion Model, HMM= Hidden Markov Model).

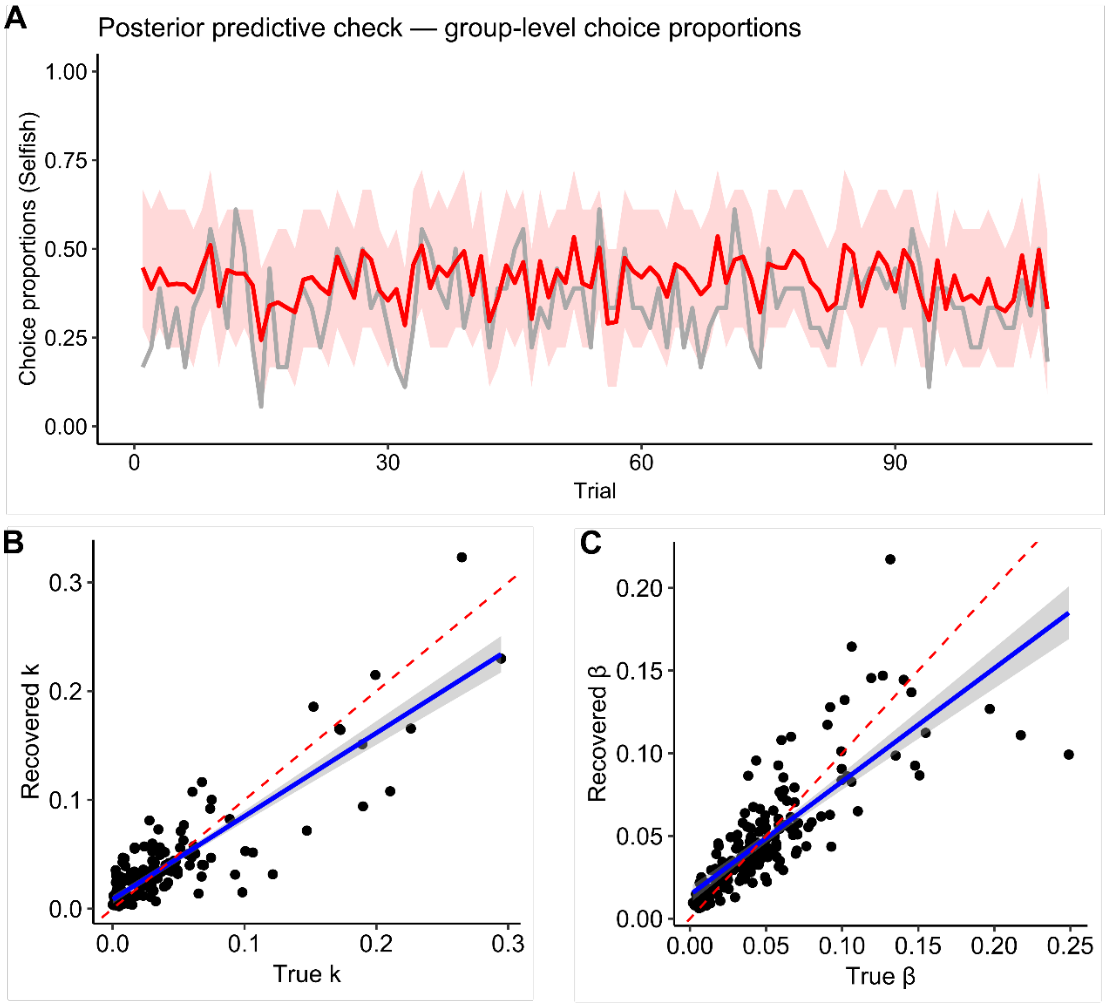


**Supp. Figure 7. Validation of the *SM Discount Model*. A**. Model predicted vs. observed choice proportions over the trial sequence. Grey line represents the empirical choices. The red line represents the most predicted choice averaged over 8000 simulated datasets; the shaded area represents 95% confidence interval across simulations. **B**. Generating vs. recovered values for the discount rate $k$. **C**. Generating vs. recovered values for the inverse temperature $\beta$. In plots B and C, the red dotted line represents the identity line.

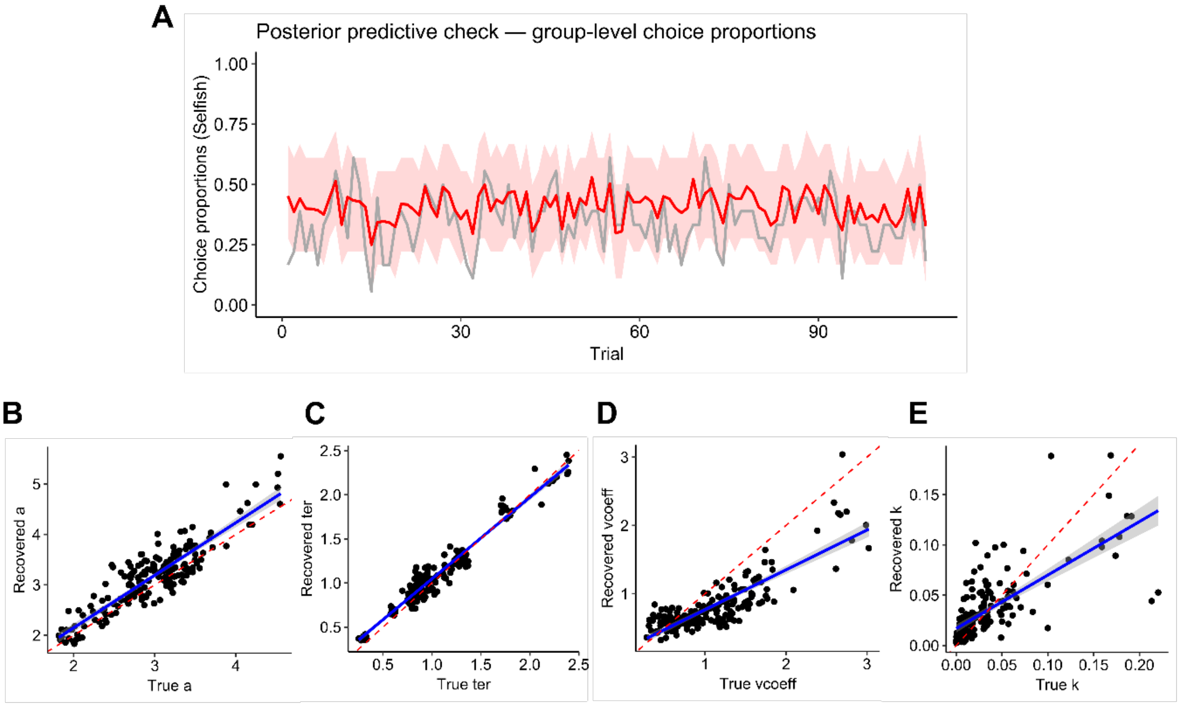


**Supp. Figure 8. Validation of the *DDM Discount Model*. A**. Model predicted vs. observed choice proportions over the trial sequence. Grey line represents the empirical choices. The red line represents the most predicted choice proportions averaged over 8000 simulated datasets; the shaded area represents 95% confidence interval across simulations. **B**. Generating vs. recovered values for the boundary separation $a$. **C**. Generating vs. recovered values for the non-decision time $ter$. **D**. Generating vs. recovered values for the drift rate coefficient $v_{coeff}$. **E**. Generating vs. recovered values for the discount rate $k$. In plots B, C, D and E, the red dotted line represents the identity line.

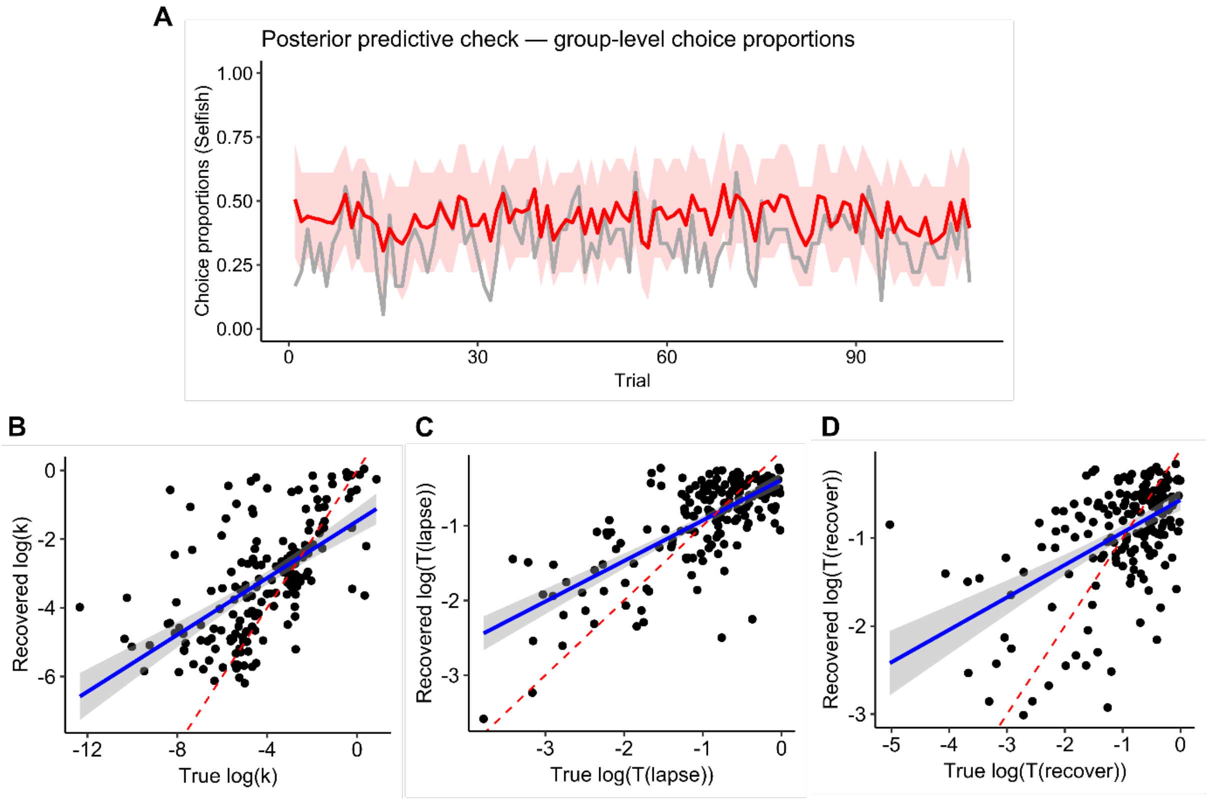


**Supp. Figure 9. Validation of the *HMM Discount Model*. A**. Model predicted vs. observed choice proportions over the trial sequence. Grey line represents the empirical choices. The red line represents the most predicted choice proportions averaged over 8000 simulated datasets; the shaded area represents 95% confidence interval across simulations. **B**. Generating vs. recovered values for r the discount rate $\log(k)$. **C**. Generating vs. recovered values for the lapse probability $\log(T_{lapse})$. **D**. Generating vs. recovered values for the recovery probability $\log(T_{recover})$. In plots B, C and D, the red dotted line represents the identity line.

#### ***Section 5***

##### Inference on posterior hyper-parameters

<u>Posterior comparisons for the *SM Discount* model</u>

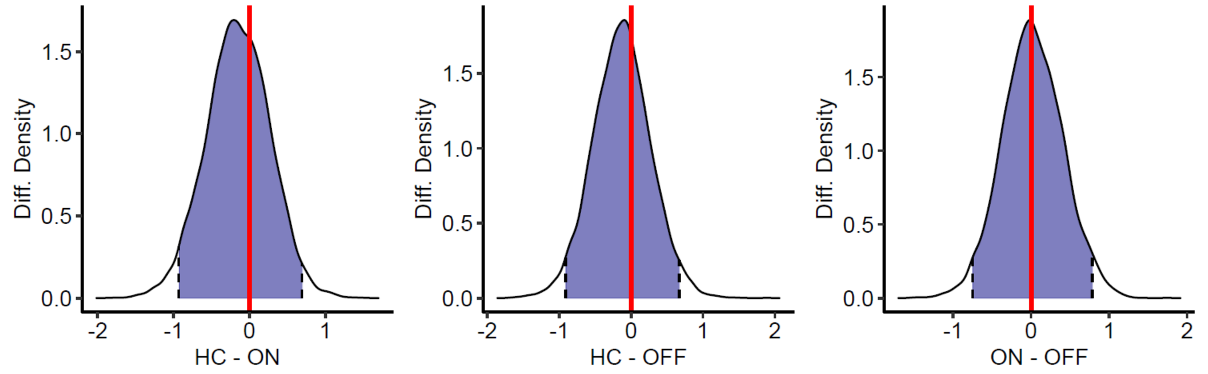


**Supp. Figure 1. Posterior comparisons for the discount rate $k$.** None of the comparisons were significant.

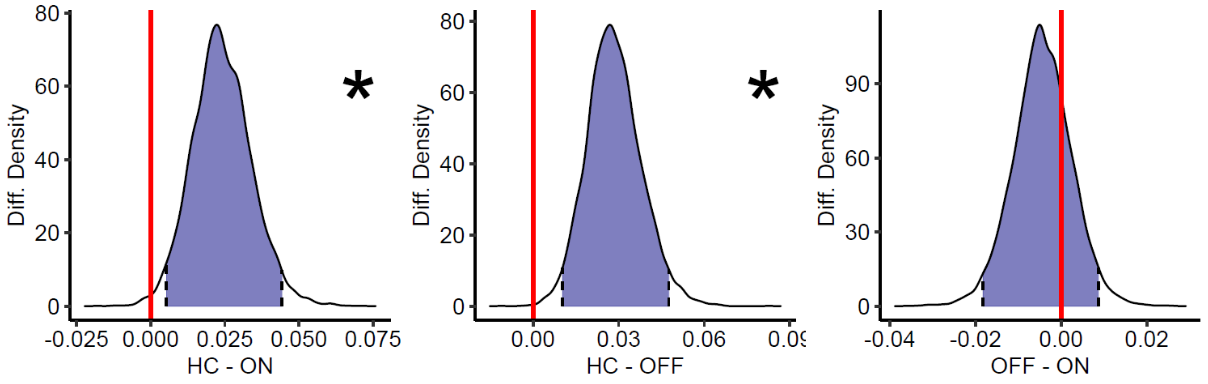


**Supp. Figure 2. Posterior comparisons for the inverse temperature $\beta$.** Significant differences are indicated with "*".

<u>Posterior comparisons for the *DDM Discount* model</u>

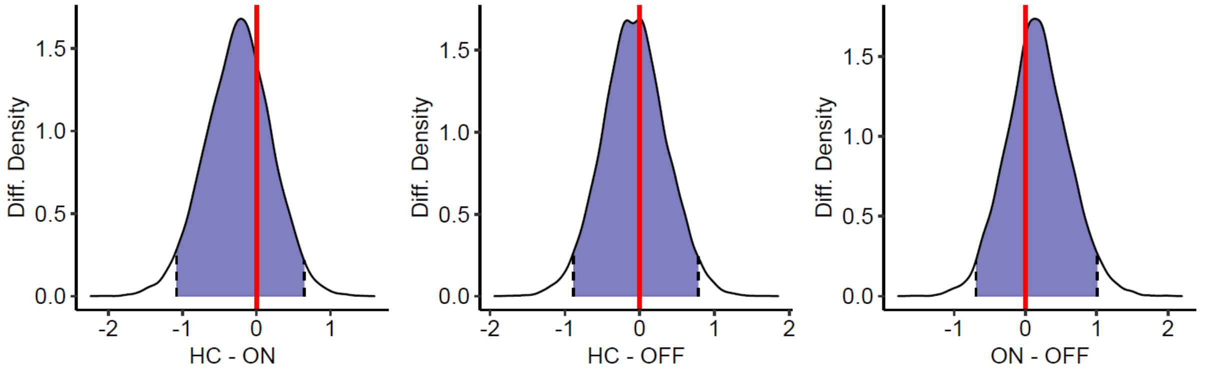


**Supp. Figure 3. Posterior comparisons for the discount rate $k$.** None of the comparisons were significant.

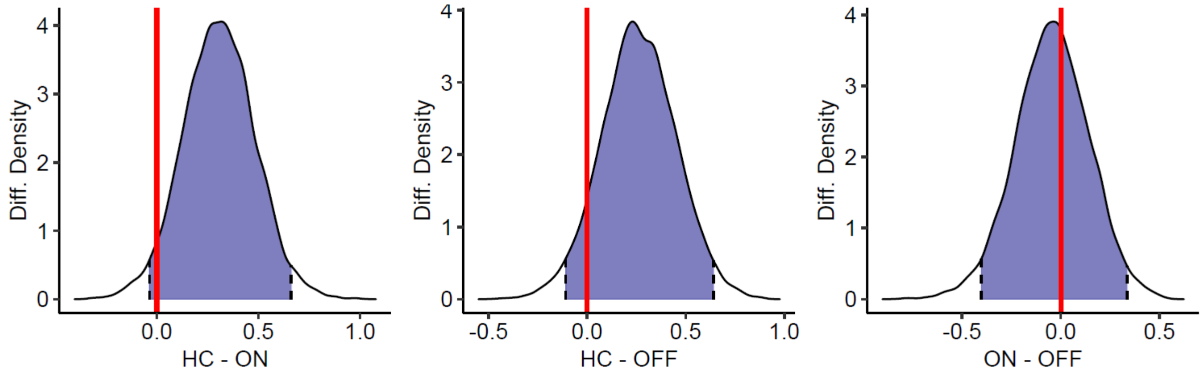


**Supp. Figure 4. Posterior comparisons for the boundary separations $a$.** None of the comparisons were significant.

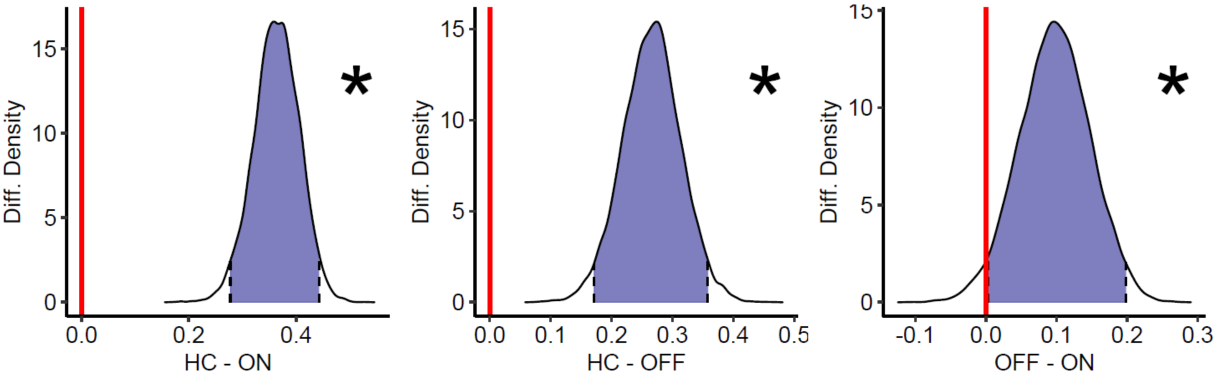


**Supp. Figure 5. Posterior comparisons for the non-decision time $ter$.** Significant differences are indicated with "*".

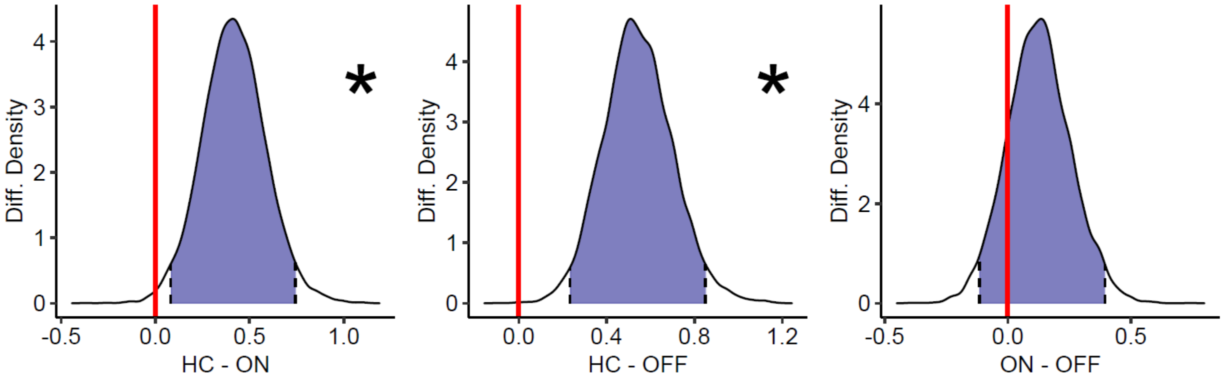


**Supp. Figure 6. Posterior comparisons for the drift rate coefficient $v_{coeff}$.** Significant differences are indicated with "*".

#### ***Section 6***

##### Correlations between electrode localizations and model parameters

In order to assess if localization of the DBS electrode within the STN affected the modelled mechanisms of social decision-making, we correlated the distance from the motor and associative sweet spots with the changes in individual-level parameters attributable to DBS stimulation, i.e. a proxy for how an individual-level parameter has changed from turning ON STN-DBS. This was done to test for possible differential effects of DBS on specific decision-making processes depending on electrode placement.

For all patients, DBS electrode localization was carried out using Lead-DBS (*v3.0*; Horn & Kühn, 2015), following established protocols, described elsewhere (Horn et al., 2019; Neudorfer et al., 2023). In brief, preoperative MRI- and postoperative CT-scans were non-linearly warped to ICBM 2009b Nonlinear Asymmetric ('MNI') space using Advanced Normalization Tools (ANTs, https://stnava.github.io/ANTs/) after co-registration. To account for potential pneumocephalus, we applied a brain shift correction. Next, the resulting transformation was further adjusted with WarpDrive, with special attention to accurate alignment of the STN (Oxenford et al., 2024). Electrode trajectories were then reconstructed using either PaCER (Husch et al., 2018) when postoperative CT data were available or TRAC/CORE (Horn & Kühn, 2015) in the case of postoperative MRI. Finally, we quantified the Euclidean proximities between the chronically active contacts and the MNI coordinates of the center of mass of both, motor ([±10.4, –11.7, –7.6 mm]) as well as associative ([±12.6, –15.0, –7.1 mm]) parts of the STN (Jorge et al., 2022). We decided to compute an estimate of anatomical proximities to both of these coordinates, since the motor STN represented the surgical target in this cohort, while associative parts of the STN might be relevant for the non-motor/cognitive DBS effects of interest in this paradigm (Schoenwald et al., 2025).

Among all possible correlations between distances to associative and motor sweet spots of the STN and changes in individual-level parameter values, there was no significant correlation.

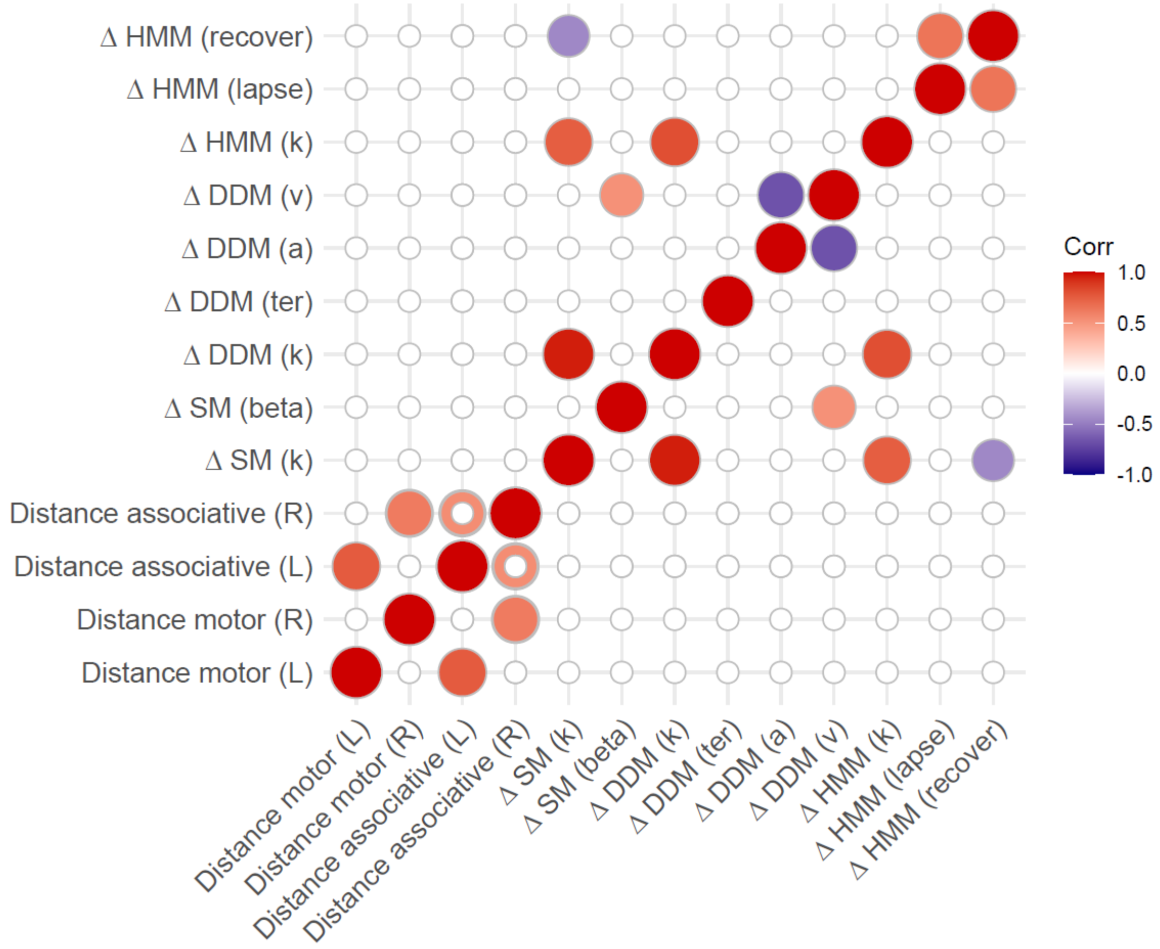


**Supp. Figure 10. Correlogram of electrode localisations and model parameters.** Shows the linear correlation between the distances from the DBS electrode to the motor and associative sweet spots of the STN and changes in model parameters attributed to stimulation, i.e. the change in parameter value from the stimulation ON to the stimulation OFF state. Only significant correlations are shown. None of the correlations between the electrode characteristics and model parameters were significant.

#### *Supplemental references*


Bates, D., Mächler, M., Bolker, B., & Walker, S. (2015). Fitting Linear Mixed-Effects Models Using lme4. *Journal of Statistical Software*, *67*, 1–48. https://doi.org/10.18637/jss.v067.i01

Carpenter, B., Gelman, A., Hoffman, M. D., Lee, D., Goodrich, B., Betancourt, M., Brubaker, M., Guo, J., Li, P., & Riddell, A. (2017). Stan: A Probabilistic Programming Language. *Journal of Statistical Software*, *76*, 1–32. https://doi.org/10.18637/jss.v076.i01

Crockett, M. J., Siegel, J. Z., Kurth-Nelson, Z., Ousdal, O. T., Story, G., Frieband, C., Grosse-Rueskamp, J. M., Dayan, P., & Dolan, R. J. (2015). Dissociable Effects of Serotonin and Dopamine on the Valuation of Harm in Moral Decision Making. *Current Biology: CB*, *25*(14), 1852–1859. https://doi.org/10.1016/j.cub.2015.05.021

Fontanesi, L., Gluth, S., Spektor, M. S., & Rieskamp, J. (2019). A reinforcement learning diffusion decision model for value-based decisions. *Psychonomic Bulletin & Review*, *26*(4), 1099–1121. https://doi.org/10.3758/s13423-018-1554-2

Frank, M. J., Samanta, J., Moustafa, A. A., & Sherman, S. J. (2007). Hold your horses: Impulsivity, deep brain stimulation, and medication in parkinsonism. *Science (New York, N.Y.)*, *318*(5854), 1309–1312. https://doi.org/10.1126/science.1146157

Horn, A., & Kühn, A. A. (2015). Lead-DBS: A toolbox for deep brain stimulation electrode localizations and visualizations. *NeuroImage*, *107*, 127–135. https://doi.org/10.1016/j.neuroimage.2014.12.002

Horn, A., Li, N., Dembek, T. A., Kappel, A., Boulay, C., Ewert, S., Tietze, A., Husch, A., Perera, T., Neumann, W.-J., Reisert, M., Si, H., Oostenveld, R., Rorden, C., Yeh, F.-C., Fang, Q., Herrington, T. M., Vorwerk, J., & Kühn, A. A. (2019). Lead-Dbs V2: Towards a Comprehensive Pipeline for Deep Brain Stimulation Imaging. *NeuroImage*, *184*, 293–316. https://doi.org/10.1016/j.neuroimage.2018.08.068

Husch, A., V. Petersen, M., Gemmar, P., Goncalves, J., & Hertel, F. (2018). PaCER - A fully automated method for electrode trajectory and contact reconstruction in deep brain stimulation. *NeuroImage: Clinical*, *17*, 80–89. https://doi.org/10.1016/J.NICL.2017.10.004

Jorge, A., Lipski, W. J., Wang, D., Crammond, D. J., Turner, R. S., & Richardson, R. M. (2022). Hyperdirect connectivity of opercular speech network to the subthalamic nucleus. *Cell Reports*, *38*(10), 110477. https://doi.org/10.1016/J.CELREP.2022.110477

Kuznetsova, A., Brockhoff, P. B., & Christensen, R. H. B. (2017). lmerTest Package: Tests in Linear Mixed Effects Models. *Journal of Statistical Software*, *82*, 1–26. https://doi.org/10.18637/jss.v082.i13

Mancini, F., Mahajan, P., Guttesen, A. á V., Onysk, J., Scholtes, I., Shenker, N., Lee, M., & Seymour, B. (2025). Enhanced behavioural and neural sensitivity to punishments in chronic pain and fatigue. *Brain*, *148*(6), 2151–2162. https://doi.org/10.1093/brain/awae408

Neudorfer, C., Butenko, K., Oxenford, S., Rajamani, N., Achtzehn, J., Goede, L., Hollunder, B., Ríos, A. S., Hart, L., Tasserie, J., Fernando, K. B., Nguyen, T. A. K., Al-Fatly, B., Vissani, M., Fox, M., Richardson, R. M., van Rienen, U., Kühn, A. A., Husch, A. D., … Horn, A. (2023). Lead-DBS v3.0: Mapping deep brain stimulation effects to local anatomy and global networks. *NeuroImage*, *268*, 119862. https://doi.org/10.1016/J.NEUROIMAGE.2023.119862

Oxenford, S., Ríos, A. S., Hollunder, B., Neudorfer, C., Boutet, A., Elias, G. J. B., Germann, J., Loh, A., Deeb, W., Salvato, B., Almeida, L., Foote, K. D., Amaral, R., Rosenberg, P. B., Tang-Wai, D. F., Wolk, D. A., Burke, A. D., Sabbagh, M. N., Salloway, S., … Horn, A. (2024). WarpDrive: Improving spatial normalization using manual refinements. *Medical Image Analysis*, *91*, 103041. https://doi.org/10.1016/J.MEDIA.2023.103041

Palminteri, S., Wyart, V., & Koechlin, E. (2017). The Importance of Falsification in Computational Cognitive Modeling. *Trends in Cognitive Sciences*, *21*(6), 425–433. https://doi.org/10.1016/j.tics.2017.03.011

Schoenwald, H., Bahners, B. H., Kannenberg, S., Dembek, T. A., Barbe, M. T., Sylaj, D., Spiewok, A., Elben, S., Muettel, T., Vesper, J., Slotty, P., Schnitzler, A., & Groiss, S. J. (2025). Antero-Lateral Subthalamic Nucleus Theta Stimulation Improves Verbal Fluency in Parkinson's Disease. *Movement Disorders*, *40*(6), 1195–1199. https://doi.org/10.1002/mds.30185

Schultz, W., Dayan, P., & Montague, P. R. (1997). A Neural Substrate of Prediction and Reward. *Science*, *275*(5306), 1593–1599. https://doi.org/10.1126/science.275.5306.1593

Wilson, R. C., & Collins, A. G. (2019). Ten simple rules for the computational modeling of behavioral data. *eLife*, *8*, e49547. https://doi.org/10.7554/eLife.49547
